# Hospital vulnerability to spread of respiratory infections: close contact data collection and mathematical modelling

**DOI:** 10.1101/2022.09.13.22279837

**Authors:** George Shirreff, Bich-Tram Huynh, Audrey Duval, Lara Cristina Pereira, Djillali Annane, Aurélien Dinh, Olivier Lambotte, Sophie Bulifon, Magali Guichardon, Sebastien Beaune, Julie Toubiana, Elsa Kermorvant-Duchemin, Gerard Chéron, Hugues Cordel, Laurent Argaud, Marion Douplat, Paul Abraham, Karim Tazarourte, Géraldine Martin-Gaujard, Philippe Vanhems, Delphine Hilliquin, Duc Nguyen, Guillaume Chelius, Antoine Fraboulet, EMEA-MESuRS Working Group on Nosocomial SARS-CoV-2 Modelling, Laura Temime, Lulla Opatowski, Didier Guillemot

**Author notes:** Equal first authorship (George Shirreff, Bich-Tram Huynh). Equal last authorship (Laura Temime, Lulla Opatowski, Didier Guillemot). Corresponding author: George Shirreff, Institut Pasteur, 25-28 rue du Dr Roux, 75015 Paris, France.

## Abstract

The transmission risk of SARS-CoV-2 within hospitals can exceed that in the general community because of more frequent close proximity interactions. However, epidemic risk across wards is still poorly described. We measured CPIs directly using wearable sensors given to all those present in a clinical ward over a 36-hour period, across 15 wards in three hospitals in spring 2020. Data were collected from 2114 participants. These data were combined with a simple transmission model describing the arrival of a single index case to the ward to estimate the risk of an outbreak. Estimated epidemic risk ranged four-fold, from 0.12 secondary infections per day in an adult emergency to 0.49 per day in general paediatrics. The risk presented by an index case in a patient varied twenty-fold across wards. Using simulation, we assessed the potential impact on outbreak risk of targeting the most connected individuals for prevention. We found that targeting those with the highest cumulative contact hours was most impactful (20% reduction for 5% of the population targeted), and on average resources were better spent targeting patients. This study reveals patterns of interactions between individuals in hospital during a pandemic and opens new routes for research into airborne nosocomial risk.

## Introduction

Hospitals are vulnerable to outbreaks of disease, which is especially important in a crisis such as the COVID-19 pandemic. During the first pandemic wave in the UK, up to 16% of COVID-19 in-patients ^1^ and 70% of staff ^2^ had acquired their infection in hospital. In addition to the direct medical risks to healthcare workers (HCW) and patients, infections among staff can lead to staff shortages and disorganisation when they are ill or forced to isolate.

A key component of infection risk for an airborne infection is the rate of close contact between individuals. This may be much higher in hospitals than in the general population, potentially leading to elevated risk of transmission ^3^. Hence, anticipating the epidemic risk and prioritising prevention measures requires an understanding of patterns of close contacts in these settings ^4^. These patterns may vary widely depending on level of activity, specialty and organisation, and indeed the proportion infected in SARS-CoV-2 outbreaks differed considerably between wards ^1,5^.

Direct recording of close proximity interactions using wearable electronic sensors enables all contacts to be recorded without inaccuracies in recall to which self-report methods are vulnerable ^6^. A limited number of previous studies have used wearable sensor technology to study interactions in hospitals. Some have relied on sensors worn only by HCWs, interacting with each other ^7^ or with fixed-point sensors which interact with the sensors worn by HCWs ^8,9^. Before the COVID-19 pandemic, studies using sensors worn by patients and HCWs have been conducted in paediatrics ^10^, geriatry ^11^, acute care ^12^ and long-term care ^13^.

This study was conducted to understand the threat of nosocomial infection during the pandemic period, by measuring patterns of contact between individuals, predicting epidemic risk and examining how to reduce it. The objectives were to collect detailed data on the frequency and duration of contacts occurring between different types of individuals across a range of different types of wards, and use this to predict epidemic risk using a simple transmission model. This would then also allow us to evaluate prevention strategies which target the most connected individuals.

## Results

Out of 2385 participants who were offered sensors, 98 (4%) refused to participate and a further 173 (7%) did not have their data recorded due to loss of their sensor. The final sample consisted of 2114 participants (89%), including 1320 HCW, 573 patients and 221 visitors, from whom 39 850 distinct interactions were recorded. Further details on the participants are shown in Supplementary *Table S2*. The contact information allowed reconstruction of the dynamic network of contact between all individuals on the ward. The contact networks exhibit different characteristic patterns, including some that are split between two separate centres, those where contacts are evenly distributed, those where a dense centre of contacts is surrounded by a lighter connected ring, or where the entire network is centralised around a hub of HCW (Figure 1).

**Figure 1.**
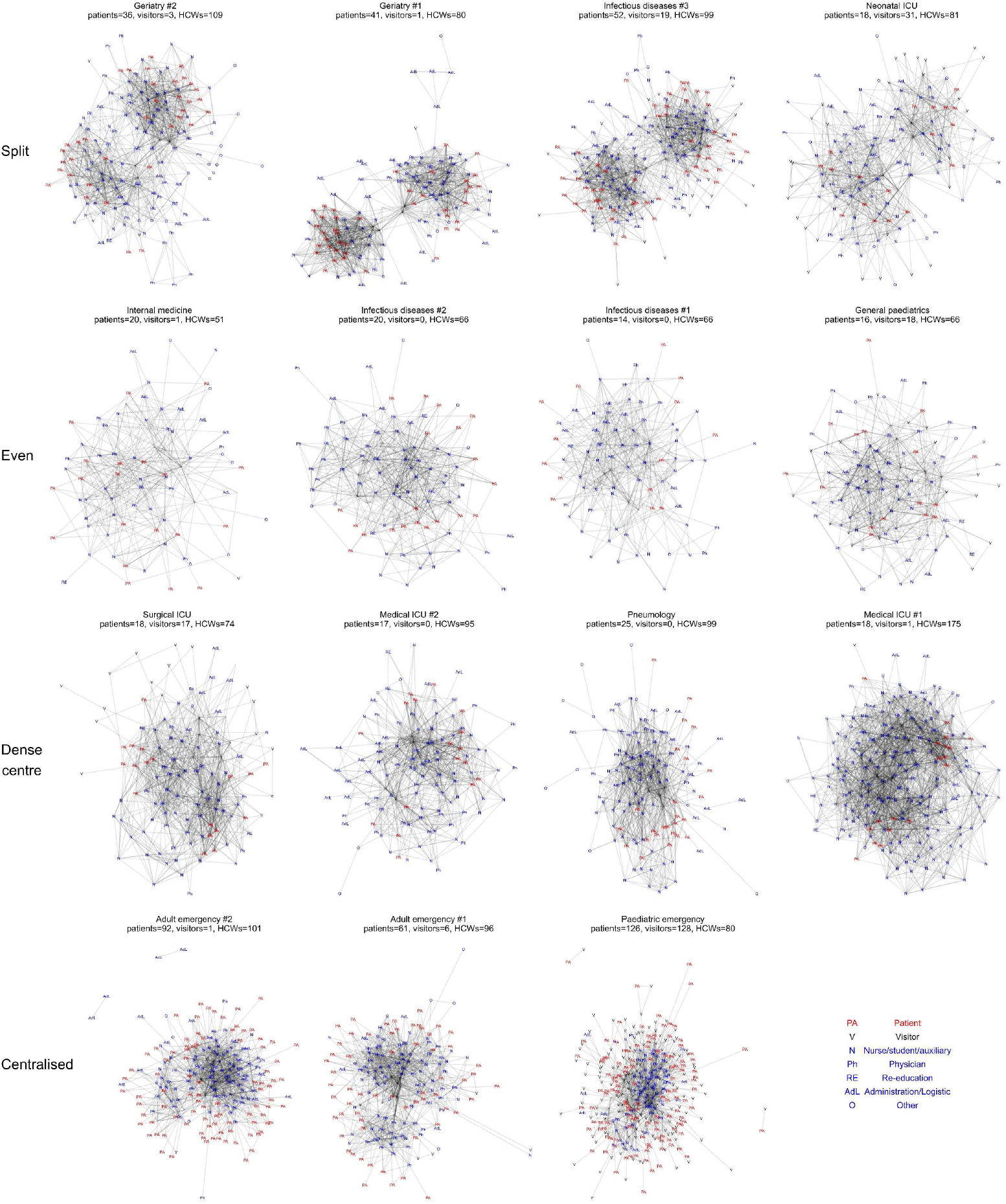
Representations of contact networks within a ward. Each individual is a node and each link a contact, regardless of duration. Each row represents a different characteristic network shape, as indicated by the labels on the left. The numbers present of each status are given in the subtitle.

### Heterogeneity in contact patterns

Contact behaviour is highly heterogeneous, as shown by the distribution of numbers of unique contacts and total contact time (Figure 2). On average, participants formed 6.7 contacts per day, with ward-level averages ranging from 4.1 to 12.5. HCW contacts are widely distributed in terms of degree while most patients have few contacts. However, in terms of total contact hours, the overall distribution is dominated by HCWs and in particular nurses and physicians.

**Figure 2.**
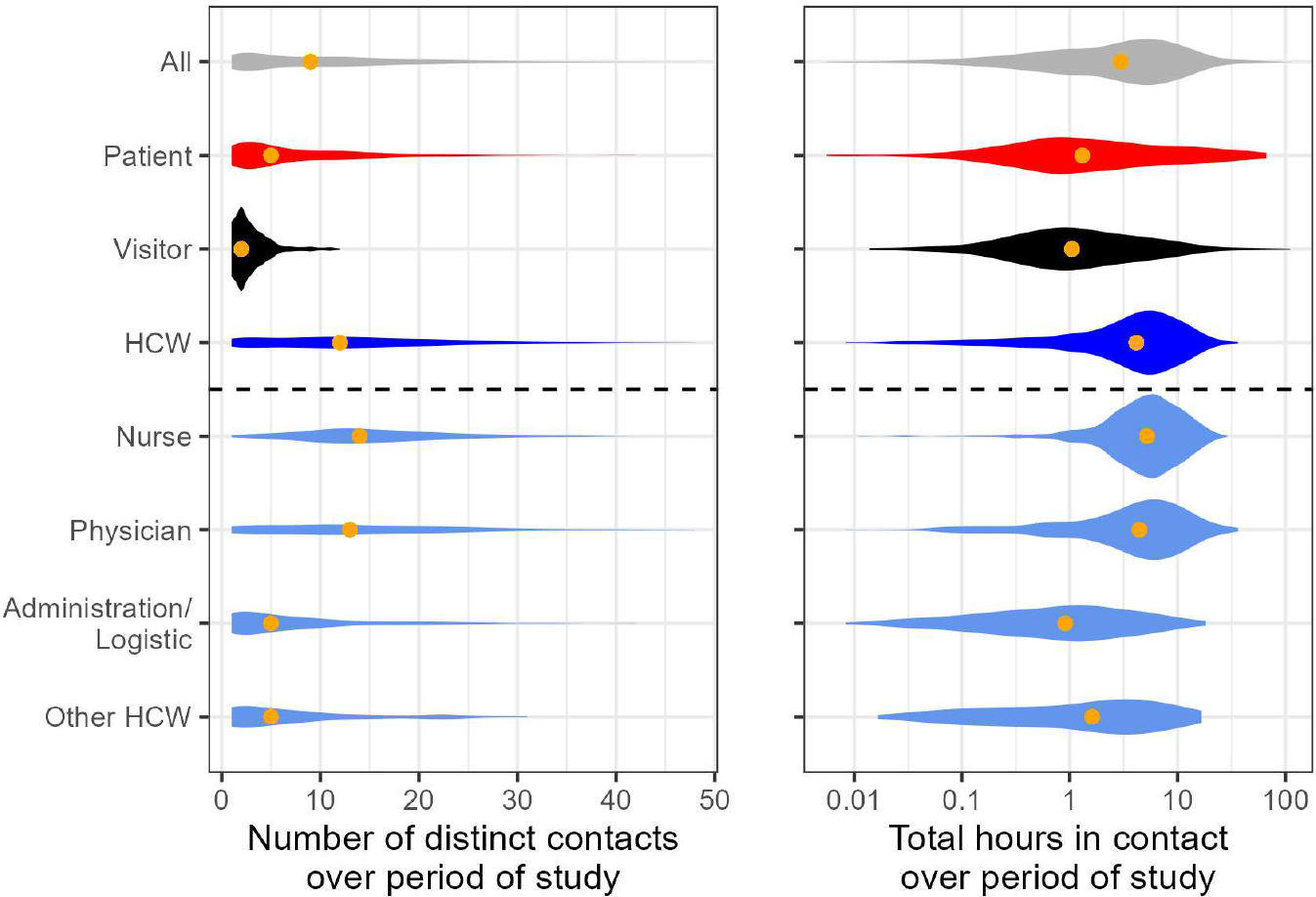
Connectivity of all status and functions of individuals across all wards. The depth of the violin represents the frequency of that value, and the total volume of each violin is equal. The orange point indicates the median of the distribution.

Average contact intensity for each status on each ward, and with every other status, is shown as a contact matrix in Figure 3. Contact intensity among HCWs is relatively consistent between wards (on average between 18 and 41 contact minutes per hour spent on the ward), with most HCW contacts occurring with other HCWs, in every ward. In 8 of the studied wards including all the ICU wards and adult emergency, patients also had the majority of their contacts with HCWs, while in the general paediatrics and paediatric emergency wards they had most of their contact with visitors, and with other patients in the remaining 5. The contact rates per hour are shown in Supplementary Figure S 2, and the average duration of contact in Supplementary Figure S 3, which shows the long duration of contacts, particularly between patients. The mean contact length was 30.3 minutes, but this varied from 15.7 to 70.6 minutes between wards.

**Figure 3.**
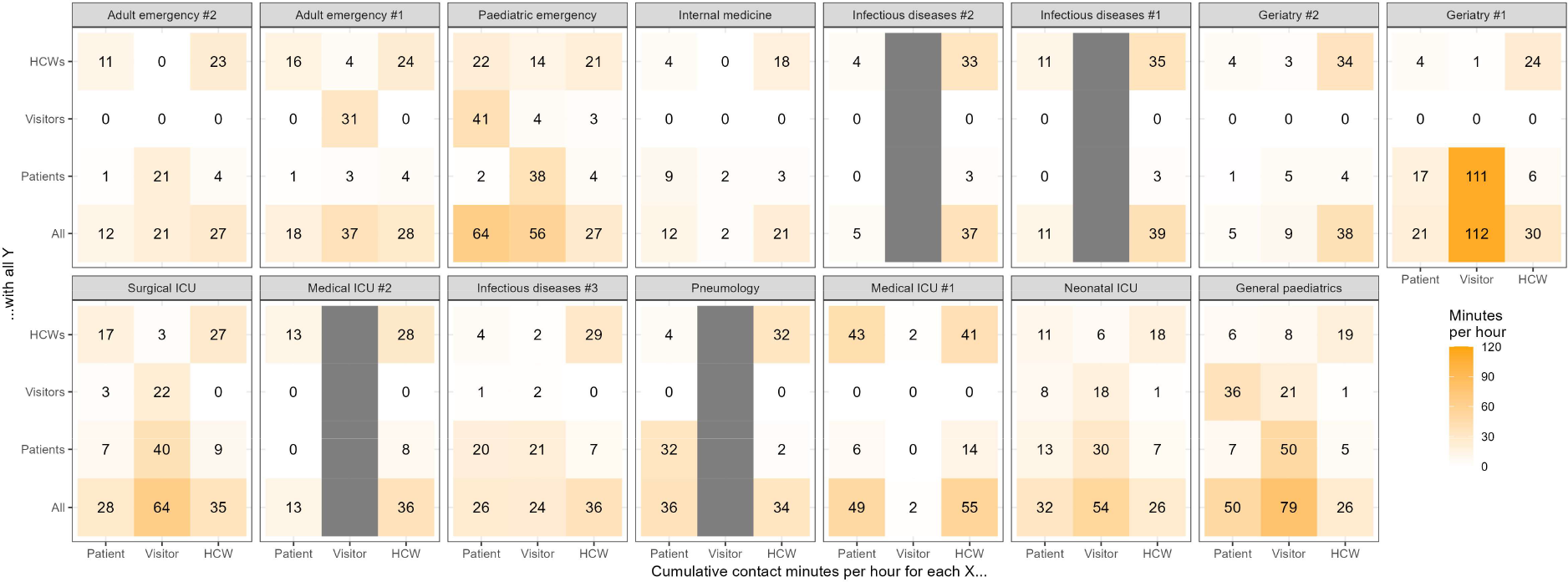
Contact intensity between statuses of individuals on each ward. Each panel represents a ward, and each cell represents the total cumulative contact minutes that each type of individual (patient, visitor or HCW, columns) has with each type of individual (rows) per hour spent carrying the sensor. Where the type of individual is not present, the corresponding column is grey.

### Variety in epidemic risk

These heterogenous contact patterns translated into heterogenous risks of an airborne pathogen spreading within the wards. Figure 4 shows that the predicted overall number of secondary infections per day varies 4-fold between the different wards, from 0.12 to 0.49, with the lowest epidemic risk in the emergency wards and highest in general paediatrics. This variation between wards is even more striking for secondary infections arising from an index case in a patient, with a predicted range from 0.04 to 0.81. In emergency units (adult and paediatric), we estimate that transmission between HCWs contributes almost all of the epidemic risk (Supplementary Figure S 4). For other wards, risk of transmission from patients was highly variable. In geriatry #1, the risk of direct patient-to-patient transmission was particularly high, while in geriatry #2 it was much lower, as in this ward the cumulative contact time between patients was considerably lower (Figure 3).

**Figure 4.**
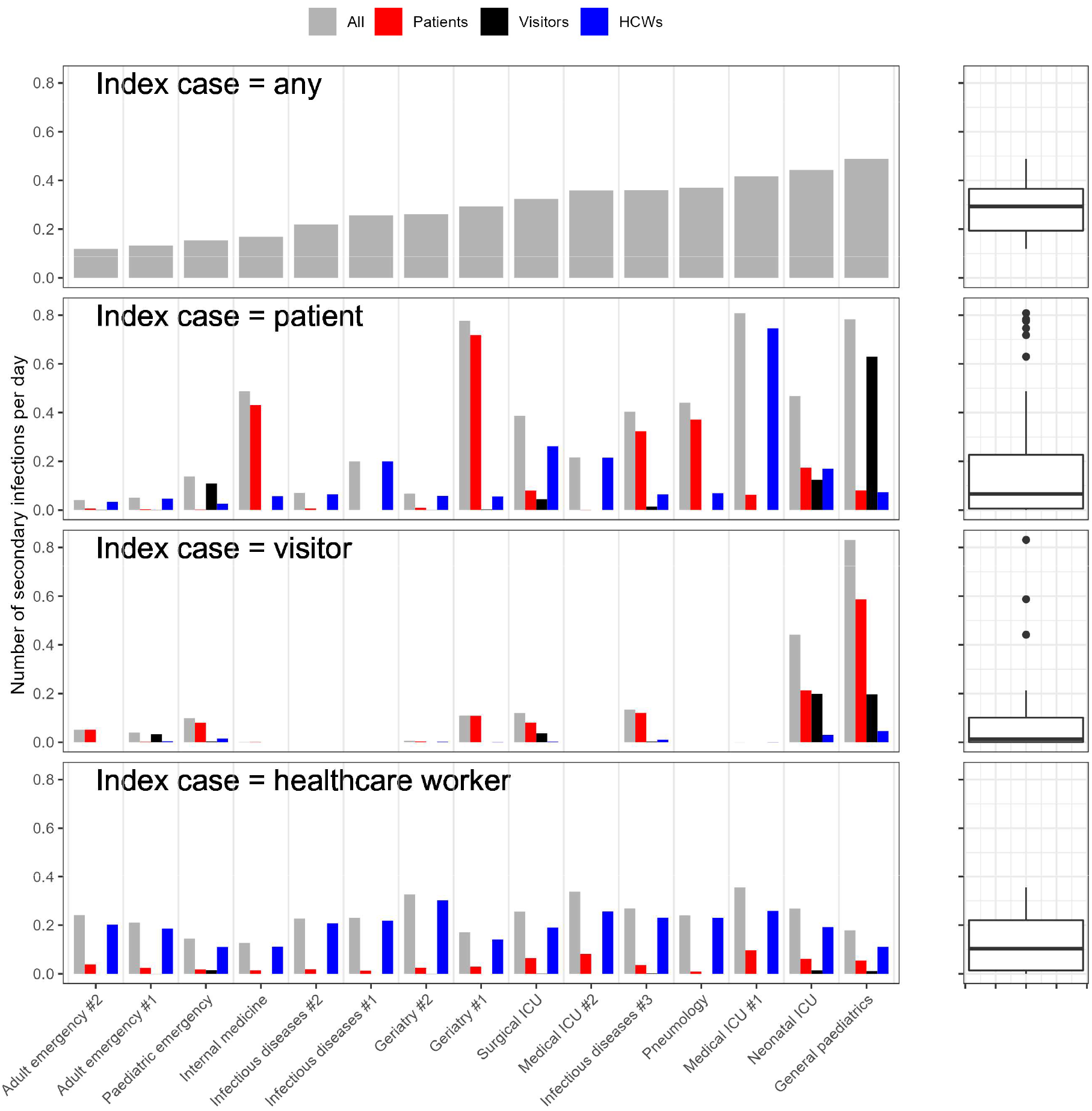
Predicted number of secondary infections per day from a single index case. Each panel represents a different hypothetical index infection, and the coloured bars represent the number of individuals of each status expected to be directly infected per day. The boxplots on the right illustrate the range of values in each bar plot.

In adult general wards, visitors presented low risk (up to 0.13 secondary infections per day). By oppositionestimated transmission risk from visitors could reach high levels in paediatric wards (up to 0.83 secondary infections per day in general paediatrics). The risk posed by HCWs was more consistent between wards (0.13 to 0.35 secondary infections per day), with other HCWs being at most risk in every ward.

### Simulating preventive interventions

Figure 5 depicts the relative reduction in the epidemic risk obtained if the most connected 5% of the population were given complete protection. The greatest effect came when targeting individuals based on their contact hours (with a 22% reduction in secondary infections in the median ward), while targeting by degree reduced infections by 13%, and selecting at random 10%. If only high-contact patients were targeted, the reduction was similar (23%), whereas only 15% could be achieved by targeting only high-contact HCWs. Much lower reductions were possible from visitors as they always made up much less than 5% of the total population size.

**Figure 5.**
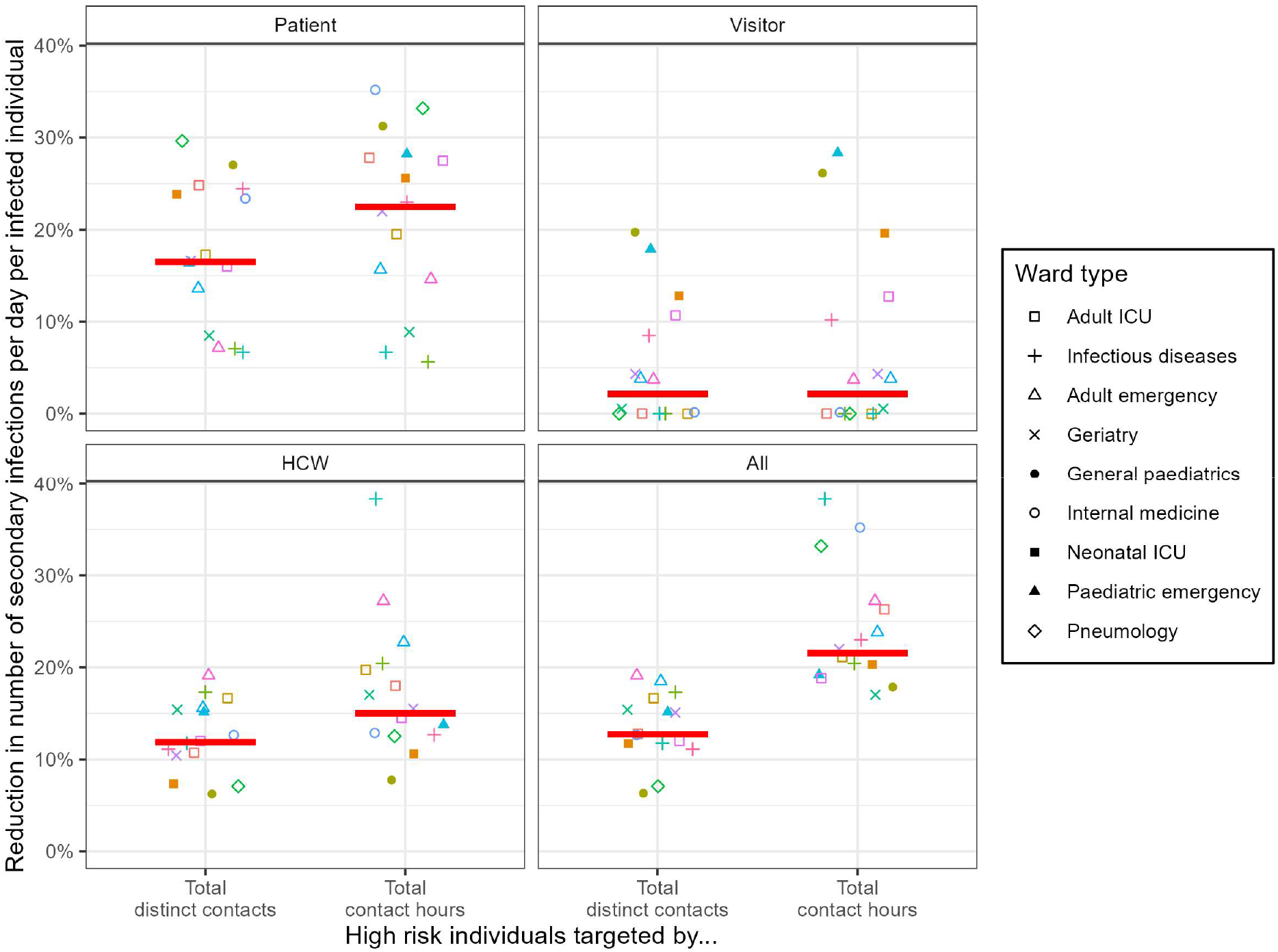
The percentage reduction in number of secondary infections per day per infected individual, when the most connected 5% of the population are completely protected. In each panel, the 5% are taken only from the indicated group. The x-axis indicates the method by which connectivity is measured for targeting. Each point represents a single ward, and the horizontal red line represents the median across all wards.

We conducted two sensitivity analyses, the first of which was to examine the effect of changing the proportion targeted over the range 0-20%. The size of the effect increased steadily with targeting, with up to a 61% reduction achievable by targeting 20% of the population (Supplementary Figure S 6). Targeting by contact hours remained the most effective method throughout, but when targeting 20% of the population, it became as effective to target HCWs as patients.

Secondly, we explored the effect of changing the shape of the relationship between time in contact and infection probability (Supplementary Figure S 5) by repeating the analysis with modified values of the shape parameter *a* (Supplementary Figure S 7). While this does change the scale of the reduction, it does not change the universal result that targeting by contact hours is the most effective. Targeting all individuals or patients was also consistently better than targeting HCWs for all values except the highest, *a*=0.5, which corresponds to a 50% chance of transmission in 2.2 hours.

## Discussion

This work reveals that the epidemic risk of an airborne pathogen, such as SARS-CoV-2, can vary widely between clinical units due to heterogeneous patterns of contacts. We find that the risk presented by a single index infection varies four-fold between wards. Emergency wards are on the lower end because the time spent by patients and visitors on the wards was too short for them to be able to transmit the virus to many others. The variation in risk rises to twenty-fold if the index case was a patient, as in some wards e.g. geriatry #1 the risk of patient-to-patient transmission was particularly high, perhaps as a result of shared activities which are typical of long-term geriatric care. Visits were generally not permitted during this period for adults, but higher risk coming from visitors was notable in paediatric wards because visitors were expressly permitted in paediatric wards, as visits were considered essential to children’s medical prognosis.

The estimated number of secondary infections reached up to 0.8 infections per day, which represents a basic reproduction number R0 of 5.6 if we assume that the index case remains infectious on the ward for 7 days ^14^. The potential for high risk implies that mandatory mask-wearing to block transmission, particularly from patients, is a valuable safety measure across all wards.

We examined how contact patterns could be exploited to improve prevention measures by better targeting, which may be critical in a context of limited resources. Our model provides an estimation of the maximum possible gain under the assumption that these measures are 100% effective, analogous to fully protective contact precautions, or complete immunisation prior to contact. As expected, targeting the most connected individuals had a disproportionate impact in reducing secondary infections. However, our work provides additional insight on how these highly-connected individuals may be selected. The biggest effect was achieved by targeting individuals by their relative contact hours. When targeting a subset of the population, the greatest overall impact was achieved by targeting patients, although the effect of targeting HCWs was more consistent. Targeting visitors was generally less effective except in paediatric wards.

The current study is, to our knowledge, the only one to have used wearable sensors to sample from all hospital users during the COVID-19 pandemic, and to have examined contact patterns across a range of specialties. A study using the same type of wearable sensors, but in a rehabilitation hospital and a pre-pandemic context, reported an average contact rate of 11.6 per day for all hospital users^13^. This estimate is higher than our own average estimate of 6.7 contacts per day, but our range of ward level averages (4.1-12.5) overlaps with this.

In our estimates of epidemic risk, we used average patterns of contact between hospital users and quantified only the risk of direct infection. While this does not take into account the dynamics of ongoing transmission across a network ^15^, we believe that our approach is more generalisable to the acute-care hospital environment, which has a largely transient population and in which therefore the connectivity between different parts of the network are less relevant.

Some limitations should be mentioned. First, the exact relationship between duration of contact and probability of infection is unknown, and is likely to differ between different SARS-CoV-2 variants. A saturating relationship between duration of exposure and infection risk has been identified, though over a timescale of days and within households ^16^. We assumed 50% probability of transmission after 11 hours of contact, but explored modifying this between 2.2 and 110 hours, which did not change our general conclusions.

Second, all types of recorded contacts were assumed to present equal risk, whereas this is likely to differ by nature of contact (e.g. conversational or physical), and be mitigated by prevention measures such as masks or hand hygiene, and vaccine-derived or natural immunity ^17^. Some care procedures may require physical contact or for the patient to be unmasked, potentially elevating the risk of patient to HCW transmission. However, since our results support the prioritisation of preventive interventions on patients over HCWs, accounting for this asymmetry should only reinforce our conclusions.

Finally, the simulations we have implemented are limited to direct short-range human-to-human transmission and do not take into account for the risk of diffusion via air flows from physically separated individuals within a clinical unit ^18,19^. However, despite the risk of longer range transmission for SARS-CoV-2, current evidence shows that droplet transmission during close proximity interactions remains key for transmission ^20^.

Beyond the illustration of its results for SARS-CoV-2, this work proposes a straightforward method based on measurements of close proximity interaction to assess and compare basic risk of airborne infection in clinical units. It allows the identification, among HCWs, patients and visitors, of those whose contribution to the global risk is highest, in order to propose priority targets for control measures. This work demonstrates the potential for combining contact monitoring and modelling to minimise nosocomial epidemic risk, which may be applied both in crisis and less urgent contexts, and adapted to other airborne bacterial or viral pathogens.

## Methods

### Data collection

The study was conducted in April-June 2020 in 15 wards in university hospital centres in Paris, Lyon and Bordeaux, selected for their diversity of clinical activity (details in Supplementary Table S 1). Each ward was studied for approximately 36 hours, starting with the nurses’ day shift in the morning of day 1 and finishing at the end of the day shift on day 2. All individuals initially present in the ward were offered sensors, as were all subsequent arrivals to the ward. At the end of the study period or on the participant’s departure, the sensor was returned. The age and function (patient, visitor, or type of health professional) of the individual was recorded, as well as the time period within which the sensor was carried. The wearable sensors (shown in Supplementary Figure S 1) recorded the identity of all other sensors within a range of about 1.5m every 10 seconds. Participants either kept the sensor in a pocket or on a pendant around the neck. For patients assigned to their room (COVID-19 patients, intensive care patients or neonates), they were hung on a fixed part of their bed.

### Contact analysis

The first step in the data analysis was to calculate summary statistics of contact, for each individual and then at the ward level between hospital users of different status (patient, visitor or HCW). The contact matrices summarise the amount of contact between each status of individual (patient, visitor and HCW) for each ward. The contact intensity and contact rate per hour, and the average duration of each contact, were calculated for individuals of status *y* with those of status *x*.

Contact intensity was calculated for each individual as the total recorded cumulative contact minutes divided by the number of hours that individual spent carrying the sensor. The contact intensity *k_xy_* is the total cumulative time an individual of status *x* spent in contact with individuals of status *y* per hour on the ward, and is calculated as in Equation [1] where *n_x_* is the number of individuals of status *x* on the ward, *i* is an individual of status *x*, *t_i_* is the number of hours this individual spent carrying the sensor, *j* is an individual of status *y*, *C_iy_* is the number of unique individuals of status *y* contacted by *i*, and *d_ij_* is the total duration of their contact over the study period.

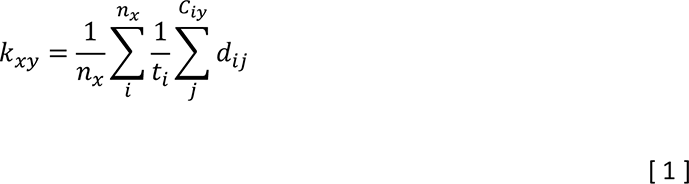

Similarly, individual contact rate was the number of unique persons contacted by that individual, per hour carrying the sensor. Average contact rate per hour *c_xy_* for individuals of status *x* with those of status *y*, is calculated by Equation [2], as the number of unique contacts of status *y* for individual *i* divided by their time with the sensor *t_i_*, and averaged over all individuals *i* of status *x*.

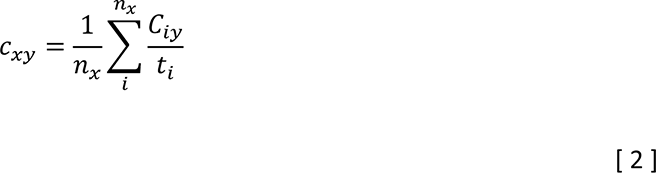

Individual average contact duration was the total cumulative contact minutes divided by the number of persons contacted. The average duration of a contact that status *x* has with status *y, d_xy_*, is calculated as in Equation [3] by first taking the average duration of all contacts an individual *i* of status *x* has with individuals *j* of status *y*, divided by all individuals of that status contacted, *C_iy_*. The average of this value is then taken across all individuals *i* of status *x*.

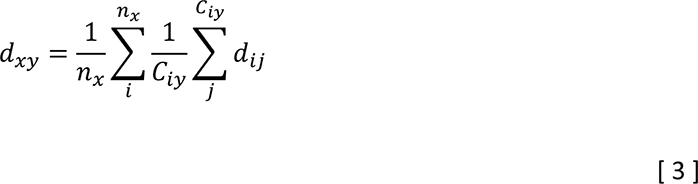

The mean of each of these measures (contact rate, contact intensity and contact duration) was then calculated for each ward and between each combination of status and provided in contact matrices.

### Epidemic risk

To examine how these ward-level values translate to epidemic risk, we wrote a transmission model to predict the number of secondary infections which would occur per day from a hypothetical SARS-CoV-2 index case if all contacts were susceptible. For each ward, we calculated the total number of expected contacts per day from the average contact rate per hour, 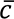 (Equation [4]) in which *n* is the total number present, *C_i_* is the total number of contacts for individual *i*.

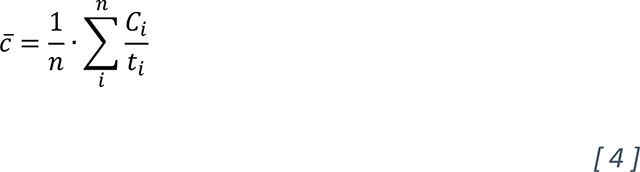

We also calculated the average time spent on the ward per 24-hour period, 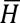 (Equation [5]), using their time carrying the sensor as a proxy, and where *T* is the total duration of the study on that ward.

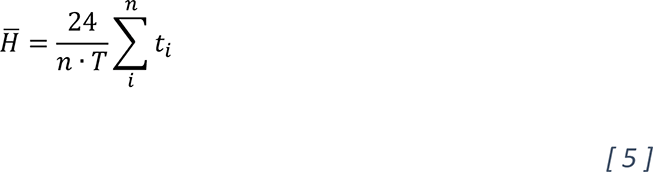

We assumed that the probability of infection per contact increased with duration of contact, and with a diminishing increase for longer contacts ^21^. The overall probability of infection per contact, 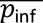 (Equation [6]), was calculated from the mean probability of infection per contact for each individual *i* across all of their contacts *j*, where the probability of infection between two individuals (Equation [7]) is determined by the duration of contact *d_ij_* and a shape parameter *a*, for which higher values are associated with a steeper increase of infection probability as contact duration increases (Supplementary Figure S 5). For the baseline analysis, a value of *a* = 0.1 is used, representing a 50% probability of infection after 11 hours in contact.

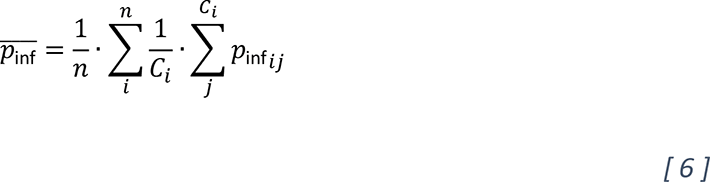

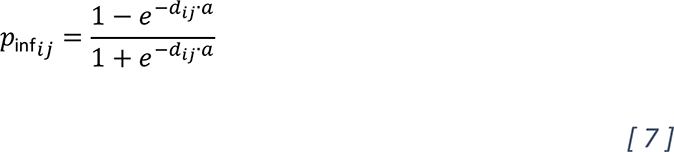

The expected number of secondary infections per day, *M*, was then computed as the product of these three quantities (Equation [8]):

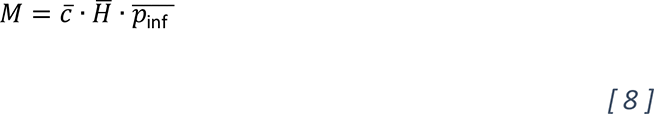

Specific predictions of numbers of secondary infections per day between different status of hospital user (patients, visitors and HCWs) were calculated using the same approach. The number of secondary infections from an index infection of status *x* towards individuals of status *y* is predicted as *M_xy_* (Equation [9]).

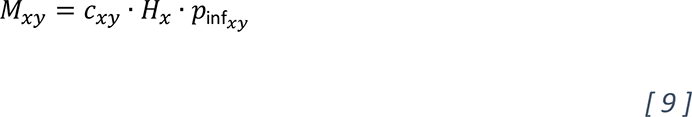

where *c_xy_* is the contact rate per hour between *x* and *y* (Equation [2]), *p*_inf_*xy*__ is the probability of infection in contacts between *x* and *y* (Equation [10], using *p*_inf_*ij*__ from Equation [7]), and *H_x_* is the average time spent on the ward by individuals of status *x* (Equation [11]).

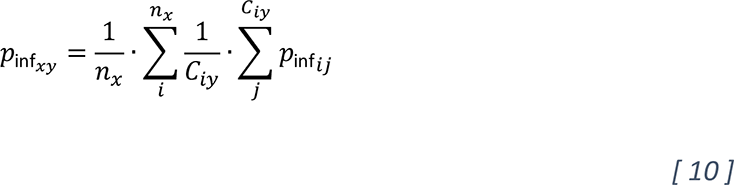

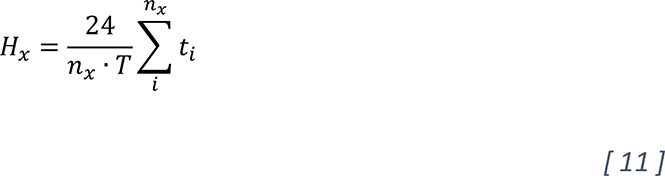

Finally, the overall number of secondary infections from an index case of status *x* to any status of individual is calculated by summing *M_xy_* over all status *y* (Equation [12]).

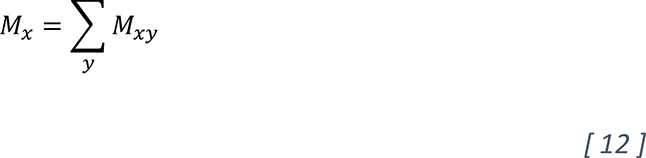

### Simulated interventions

We used this model to predict the effect of control measures targeting the most connected individuals by repeating this calculation of epidemic risk, *M*, but with the highest risk individuals being neither susceptible nor capable of transmitting. We selected the 5% of the population with either the most unique contacts over the whole study period, or the highest cumulative contact hours. The probability of infection from or to these individuals was set to zero. We also evaluated the targeting of only individuals of a single status, e.g. highly connected patients, ensuring for comparability that the number targeted still made up 5% of the total population. The reduction in daily risk was calculated as a proportion of the baseline risk in which nobody was targeted (Equation [13]).

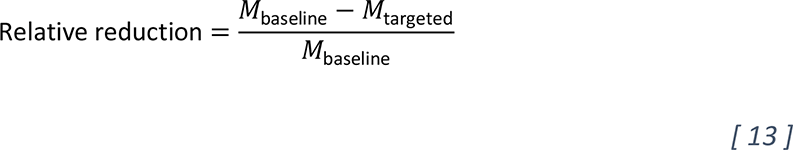

We tested the sensitivity of the simulation analysis to the proportion of the population targeted (over the range 0%-20%) and the shape parameter *a* which drives the increase in the infection probability for longer contacts (over the range 0.05-0.5).

All analyses were conducted using *R 4.2.0* ^22^, with network analyses conducted using *igraph*, and graphics produced using *ggplot2*. The code used for each analysis and visualisation is available at https://github.com/georgeshirreff/nodscov2_risksim.

### Ethics approval and consent to participate

This research was approved by the Comités de protection des personnes (CPP) Ile-de-France VI on 14/04/2020 and the Commission nationale de l’informatique et des libertés (CNIL) on 16/04/2020. Signed consent by patients, medical and administrative staff, and visitors was not required according to the CPP and CNIL, but participants could refuse to participate. When patients were minors, unable to refuse or under guardianship, parents, family or guardians, respectively, were asked. The study was carried out in accordance with the Declaration of Helsinki.

## Data Availability

All code used for analysis and visualization is available at https://github.com/georgeshirreff/nodscov2_risksim along with a subset of the data.

https://gitlab.pasteur.fr/gshirref/nodscov2_risksim

## Acknowledgements

We would like to thank: Nawal Derridj-Ait Younes, Naima Sghiouar, Tanga Vanessa Ntouba Christianne, Sylvie Azerad and Théo Debert from the clinical research unit of the Paris-Saclay university hospital; Gaetane Niel, Ga Han Park, Audrey Vallerix, Cypriane Tazi, Loueli Ouballa, Valentine Le Cardonnel, Lou Davaine, Madeleine Dutheil de la Rochère, Adeline Alleau, Tiphaine Biaggi, Manuela Carrico, Antoine Goudour, Pauline Jaubert, Marion Galliou, Mathilde de Menthon, Noémie Chanson for their participation in the field investigation; Ajmal Oodally for discussions around the network analysis; The EMEA-MESuRS working group on the nosocomial modelling of SARS-CoV-2, of whom additional members are as follows: Sophie Chervet, Kévin Jean, Sofía Jijón, David RM Smith, Ajmal Oodally, Niels Hendrickx, Cynthia Tamandjou.

## Authors’ contributions

Study conception by DG, BTH, LO, LT; data collection supervised by BTH, DG; data collection conducted by BTH, DG, DA, A Dinh, OL, SB, MG, SB, JT, EKD, GC, HC, LA, MD, PA, KT, GMG, PV, DH, DN; analyses supervised by GS, DG, BTH, LO, LT; network analysis by GS, A Duval; contact matrix analysis by GS, A Duval, LCP; epidemic risk models by GS, LO, LT; writing of original draft by GS, DG; reviewing and editing of draft by GS, DG, BTH, LO, LT. All authors have read and approved the final manuscript.

## List of abbreviations

CNIL: Commission nationale de l’informatique et des libertés
COVID-19: Coronavirus Disease 19
CPP: Comités de protection des personnes
HCW: HCWs Healthcare worker, healthcare workers
SARS-CoV-2: Severe Acute Respiratory Syndrome Coronavirus 2

