## Supplementary Material for "Hospital vulnerability to spread of respiratory infections: close contact data collection and mathematical modelling"

### Supplementary Figures

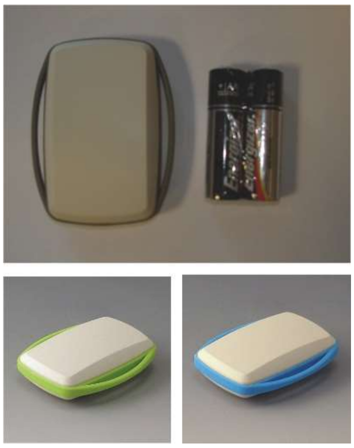

***Supplementary Figure S 1. Wearable sensor for measurement of close proximity interactions.*** *WSN430 wearable sensor, about 4cm x 6cm. Sensors are powered by a TI MSP430 micro-controller and communicates through a TI Chipcon CC1100 packet radio interface, with a memory and a battery. Sensors were kept in a low-power mode (sleep mode) for most of the time, and awakened every 10 seconds to broadcast information (a "Hello" packet). When a sensor received this from another sensor, it logged that interaction and the time. Data from the sensor was uploaded upon its return.*

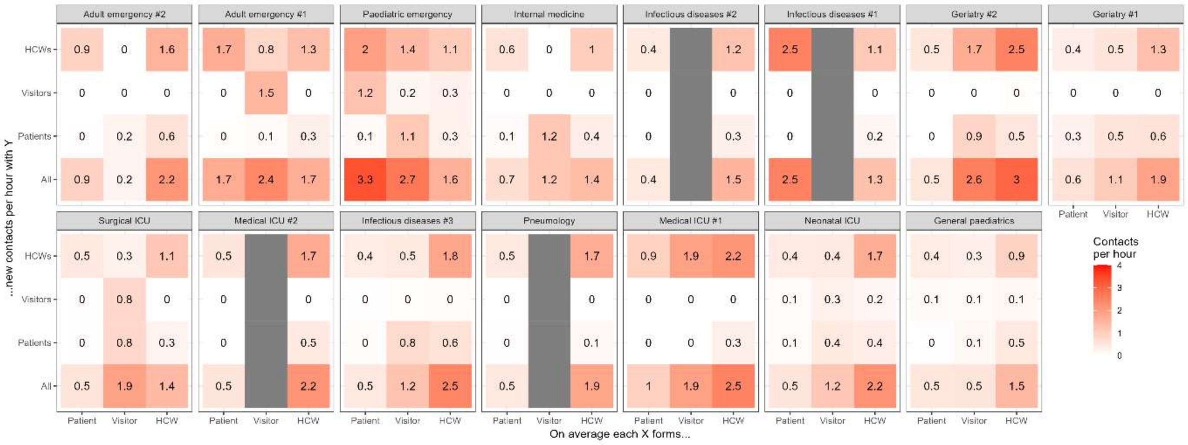


***Supplementary Figure S 2. Contact rate between types of individuals on each ward.*** *Each panel represents a ward, and each cell represents the contact rate that each status of individual (patient, visitor or HCW, columns) has with each type of individual (rows). Contact rate is the number of unique new contacts formed throughout the study period, per hour spent carrying the sensor. Where the type of individual is not present, the corresponding column is is grey.*

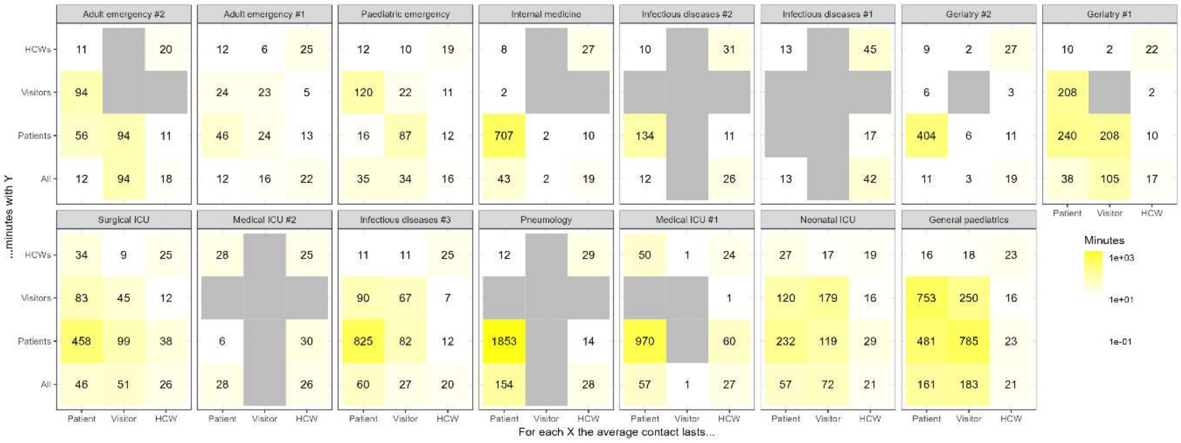


***Supplementary Figure S 3. Duration of contacts between types of individuals on each ward.*** *Each panel represents a ward, and each cell represents the average contact duration that each type of individual (patient, visitor or HCW, columns) has with an individual of each type (rows). Average contact duration is calculated for each individual with status Y, before the average for the column is calculated across all individuals of status X. Where the individual in the row and column have no contact, the average duration of contact is not calculable and the cell is grey.*

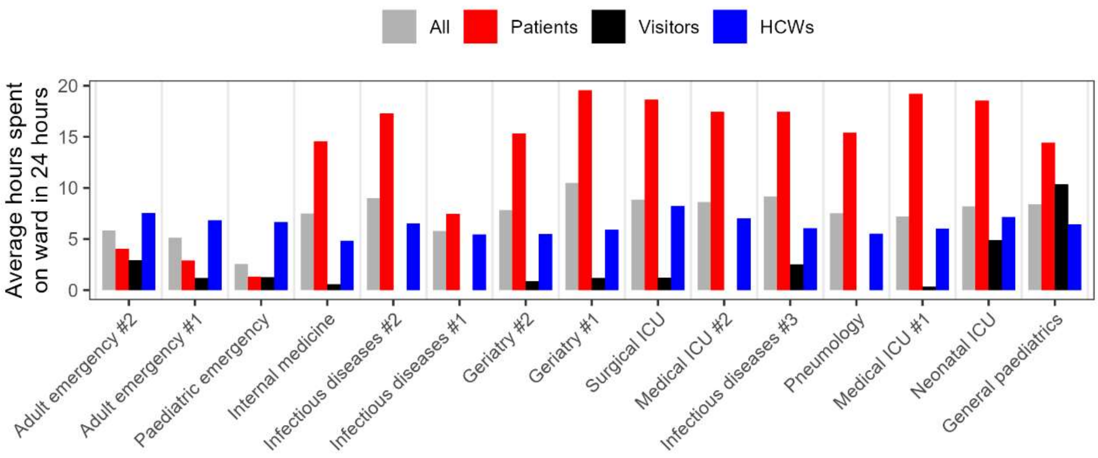


*Supplementary Figure S 4. Average amount of time spent on the ward (carrying the sensor) by each status of individual, normalised to 24 hours.*

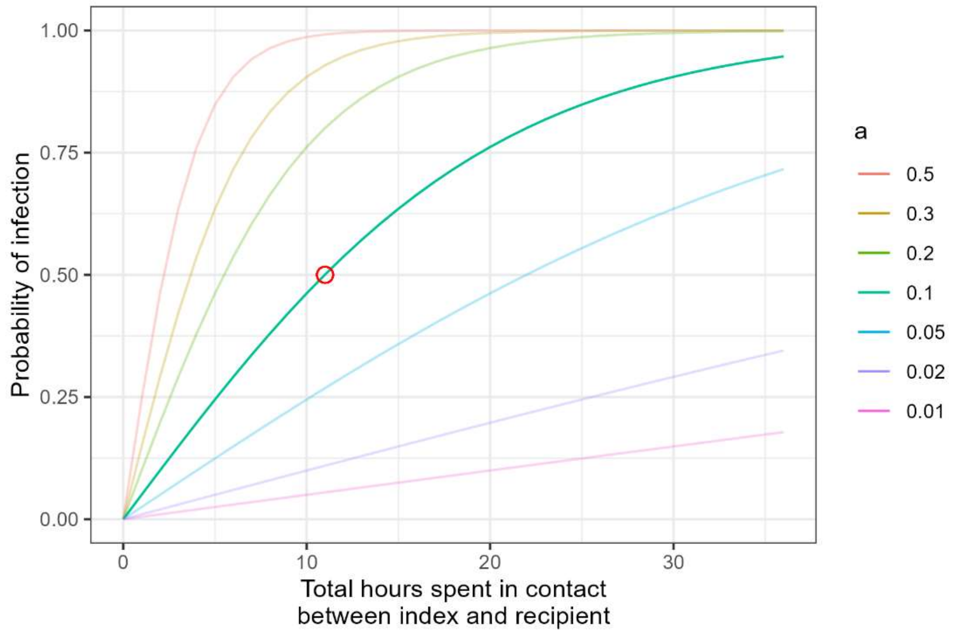


*Supplementary Figure S 5. Relationship between duration of contact and probability of infection. For different values of* a*, the probability of infection during a contact of duration hours between an infectious and susceptible individual, according to Equation [ 7 ]. The value a=0.1 is used for the baseline analysis, and for illustration, the time at which probability of infection is 50% is marked with a red circle.*

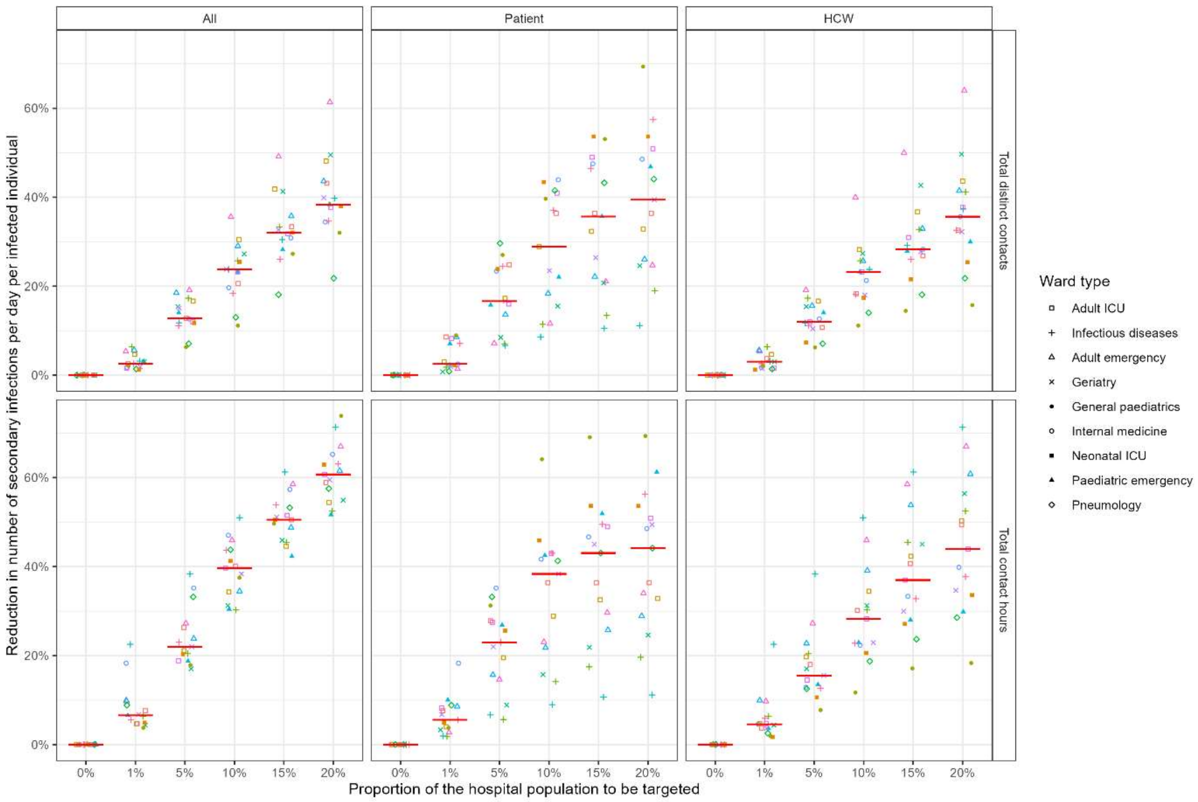


*Supplementary Figure S 6. The percentage reduction in number of secondary infections per day per infected individual when the most connected X% part of the population are completely protected. In each panel, the indicated proportion are taken only from the indicated group. Not all groups were included because often the smaller groups made up substantially fewer than 20% of the whole*

*population. The x-axis indicates the proportion protected. Each point represents a single ward, and the horizontal red line represents the median across all wards.*

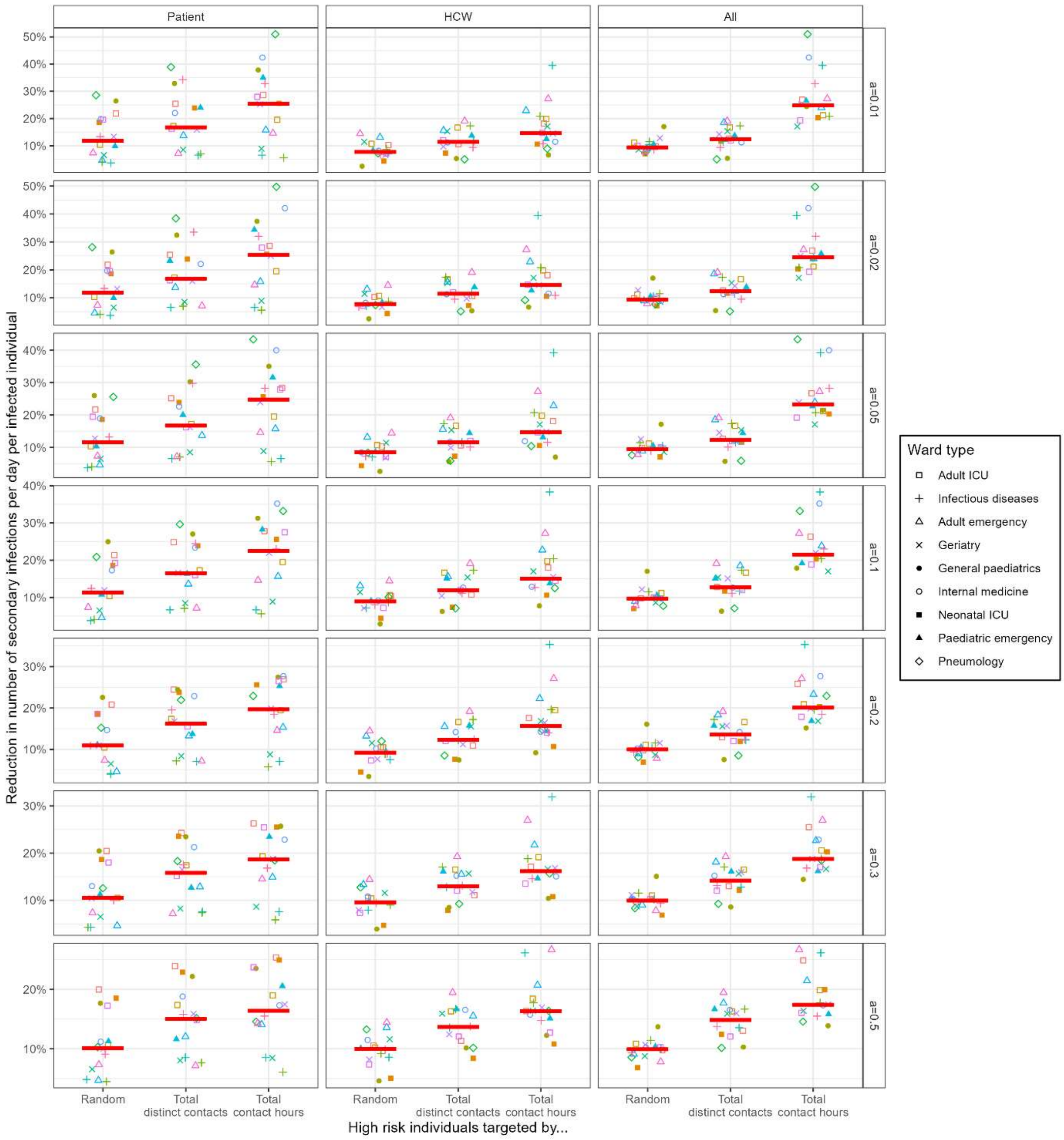


*Supplementary Figure S 7. For different values of the shape parameter* a *(rows of panels), the percentage reduction in risk is calculated when the most connected 5% of the population are completely protected. In each panel, the 5% are taken only from group indicated on the column. The x-axis indicates the targeting method. The horizontal red line indicates the median across all wards.*

### Supplementary Tables

*Supplementary Table S 1.* ***List of wards investigated.*** *Regional weekly hospitalization was the 7-day moving average of hospital admissions with SARS-CoV-2 in the corresponding region on the first day of the study (Sante Publique France, tx_indic_7J_hosp). The total hospital users refers to the number of HCW, patients and visitors who had contact data recorded during the study. The peak hospital users is the most number of these individuals who were present at any one time.*

| Medical specialty | Date of investigation | Regional weekly hospitalisation | Total hospital users | Peak hospital users |
|---|---|---|---|---|
| Infectious diseases #1 | 2020, April 17-18 | 33.12 | 80 | 45 |
| Pneumology | 2020, April 20-21 | 28.61 | 124 | 60 |
| Internal medicine | 2020, April 24-25 | 21.09 | 72 | 39 |
| Infectious diseases #2 | 2020, May 4-5 | 10.30 | 86 | 50 |
| Medical ICU #1 | 2020, May 6-7 | 9.18 | 194 | 88 |
| General paediatrics | 2020, May 13-14 | 6.35 | 100 | 53 |
| Geriatry #1 | 2020, May 18-19 | 5.32 | 122 | 82 |
| Paediatric emergency | 2020, May 21-22 | 3.96 | 334 | 64 |
| Neonatal ICU | 2020, May 25-26 | 3.22 | 130 | 69 |
| Geriatry #2 | 2020, June 4-5 | 0.76 | 148 | 84 |
| Medical ICU #2 | 2020, June 9-10 | 0.54 | 112 | 61 |
| Adult emergency #1 | 2020, June 11-12 | 0.50 | 163 | 51 |
| Surgical ICU | 2020, June 16-17 | 0.60 | 109 | 64 |
| Adult emergency #2 | 2020, June 18-19 | 0.56 | 194 | 72 |
| Infectious diseases #3 | 2020, June 23-24 | 0.07 | 170 | 98 |

*Supplementary Table S 2.* ***Population description by category.*** *Total approached, the number (and proportion) who participated, who refused, for whom the captor was lost (and data therefore unrecorded), and the mean age of participants.*

| Category | Total | Participated | Refused | Lost | Mean age (range) |
|---|---|---|---|---|---|
| Nurse | 359 | 347 (97%) | 6 (2%) | 6 (2%) | 34 (22-64) |
| Physician | 305 | 277 (91%) | 4 (1%) | 24 (8%) | 36 (24-68) |
| Auxilliary nurse | 280 | 262 (94%) | 11 (4%) | 7 (3%) | 38 (20-62) |
| Other | 124 | 105 (85%) | 2 (2%) | 17 (14%) | 39 (19-64) |
| Administratitive | 98 | 89 (91%) | 2 (2%) | 7 (7%) | 46 (23-67) |
| Student physician | 92 | 86 (93%) | 2 (2%) | 4 (4%) | 24 (21-52) |
| Logistic | 109 | 69 (63%) | 5 (5%) | 35 (32%) | 38 (19-63) |
| Student nurse | 47 | 46 (98%) | 0 | 1 (2%) | 26 (19-51) |
| Reeducation | 39 | 39 (100%) | 0 | 0 | 32 (22-57) |
| Children (0-18) | 239 | 160 (67%) | 49 (21%) | 30 (13%) | 6 (0-18) |
| Adults | 413 | 413 (100%) | 0 | 0 | 65 (19-103) |
| Visitor | 280 | 221 (79%) | 17 (6%) | 42 (15%) | 41 (0-88) |
| Total | 2385 | 2114 (89%) | 98 (4%) | 173 (7%) | 40 (0-103) |
